# The Barrier to Vaccination Is Not Vaccine Hesitancy: Patterns of COVID-19 Vaccine Acceptance over the Course of the Pandemic in 23 Countries

**DOI:** 10.1101/2021.04.23.21253857

**Authors:** Ammina Kothari, Gerit Pfuhl, David Schieferdecker, Casey Taggart Harris, Caitlin Tidwell, Kevin M Fitzpatrick, Stephanie Godleski, Saurabh Sanjay

**Affiliations:** School of Communication, Rochester Institute of Technology; Department of Psychology, UiT The Arctic University of Norway; Institute of Media and Communications, Free University Berlin, Germany; Department of Sociology and Criminal Justice, University of Arkansas; Department of Psychology, Rochester Institute of Technology; Golisano College of Computing and Information Sciences Rochester Institute of Technology

**Keywords:** COVID-19, vaccine acceptance, public health, global health, public opinion, longitudinal data, cross-country comparison

## Abstract

**Background:** At present, evidence is inconclusive regarding what factors influence vaccine intent, and whether there are widespread disparities across populations and time. The current study provides new insights regarding vaccine intent and potential differences across 23 countries and over time.

**Methods:** Our data come from a unique longitudinal survey that contains responses from Facebook users (N=1,425,172) from the 23 countries from four continents collected in 18 waves from July 2020 through March 2021.

**Results:** We find that vaccine intent varies significantly across countries and over time. Across countries, there are notable disparities in intent to vaccinate. Regarding time, intent has recently reached an all-time high. Our data demonstrates that intent to vaccinate has increased as countries have deployed vaccines on larger scales with undecidedness declining. However, there are some countries where vaccine intent is stagnant and in one country – Egypt – where it seems to have declined.

**Interpretations:** Large numbers of citizens across the world are willing to get vaccinated. In the vast majority of countries in our sample, these were high enough to reach more conservative levels of herd immunity^1^ if combined with numbers of persons already infected. As such, the main barrier to vaccination is not vaccine hesitancy, but the shortage of vaccines. This sends a clear message to politicians who need to work on a quick and fair distribution of vaccine; and to scientists who need to focus their attention on understanding remaining pockets of vaccine skepticism or undecidedness and on factors that explain actual vaccine behavior, rather than intent.

## Introduction

Hope for curbing the global spread of COVID-19 pandemic rests on immunization. That the public is willing to get vaccinated is a necessary condition for the success of this strategy. However, vaccine hesitancy remains one of the threats to global health^2^. As long as segments of the global population are hesitant to get vaccinated, new geographic variants may emerge that escape the vaccine and re-introduce the virus in vaccinated populations.

Vaccine intent is shaped by a multitude of factors, including mutual trust between communities and public health sectors^3,4,5,6^ and fact-saturated information ecologies that reduce doubts created by misinformation^7,8,9^. For example, people willing to get vaccinated has been found to be lower in countries with a history of vaccine mistrust.^13^ Likewise, common vaccination practices within a country may also impact pandemic-related vaccine acceptance in ways that reflect cultural or political disparities^10^.

For the COVID-19 pandemic, knowledge of vaccine intent remains incomplete. Yet, emerging studies suggest some variation across time and space^6-11^. For example, survey findings from earlier phases of the pandemic suggest relatively low levels of intent to vaccinate against COVID-19, particularly in the Middle East, Russia, Africa, and several European countries^12^. Other research points to an increase in vaccination intent between November 2020 and mid-January 2021 in 11 of 15 countries surveyed, but notably lower rates in countries with a history of vaccine mistrust^13^.

Based on this fragmentary evidence, we investigate the levels of vaccine acceptance over both a longer time span that covers more recent developments (e.g., emergency use authorizations, additional clinical trials, etc.), as well as across a breadth of countries to provide greater context to understanding vaccine intent. We, therefore, ask how willing are people to get the COVID-19 vaccination around the globe? Subsequently, we also ask how has this changed over the course of the pandemic? in addition to countries over time, we further explicate the geographic variation in vaccine intent within the US for better understanding meso-level patterns of vaccine skepticism. Doing so illustrates the importance of considering the unique geographic contexts in which population health outcomes are manifested. Answering these questions provides clearer understanding of whether vaccine hesitancy remains a barrier to global vaccination, and where public health practitioners and policy advocates should look to identify pockets of vaccine hesitancy.

## Methodology

### Participants and Sampling Strategy

Our data come from the global survey of Facebook users on COVID-19 beliefs, behaviors and norms implemented by the Massachusetts Institute of Technology in collaboration with Facebook, and with input from researchers at Johns Hopkins University (JHU), the World Health Organization (WHO), and the Global Outbreak Alert and Response Network (GOARN).^14^ Data were collected in two-week increments from July 2020 through March 2021, amounting to 18 survey waves. The data captures responses from individuals from 23 other countries: nine in Asia (Bangladesh, India, Indonesia, Japan, Malaysia, Pakistan, Philippines, Thailand, Vietnam), two in Africa (Egypt, Nigeria), seven in Europe (France, Germany, Italy, Poland, Romania, Turkey, United Kingdom), the United States, and four in Latin and South America (Argentina, Brazil, Colombia, Mexico). Table 1 breaks down our response rate by gender and age group from each country pooled over the 18 waves^1^ ; the technical report^13^ lists full details for the demographics of the sample.

**Table 1:**
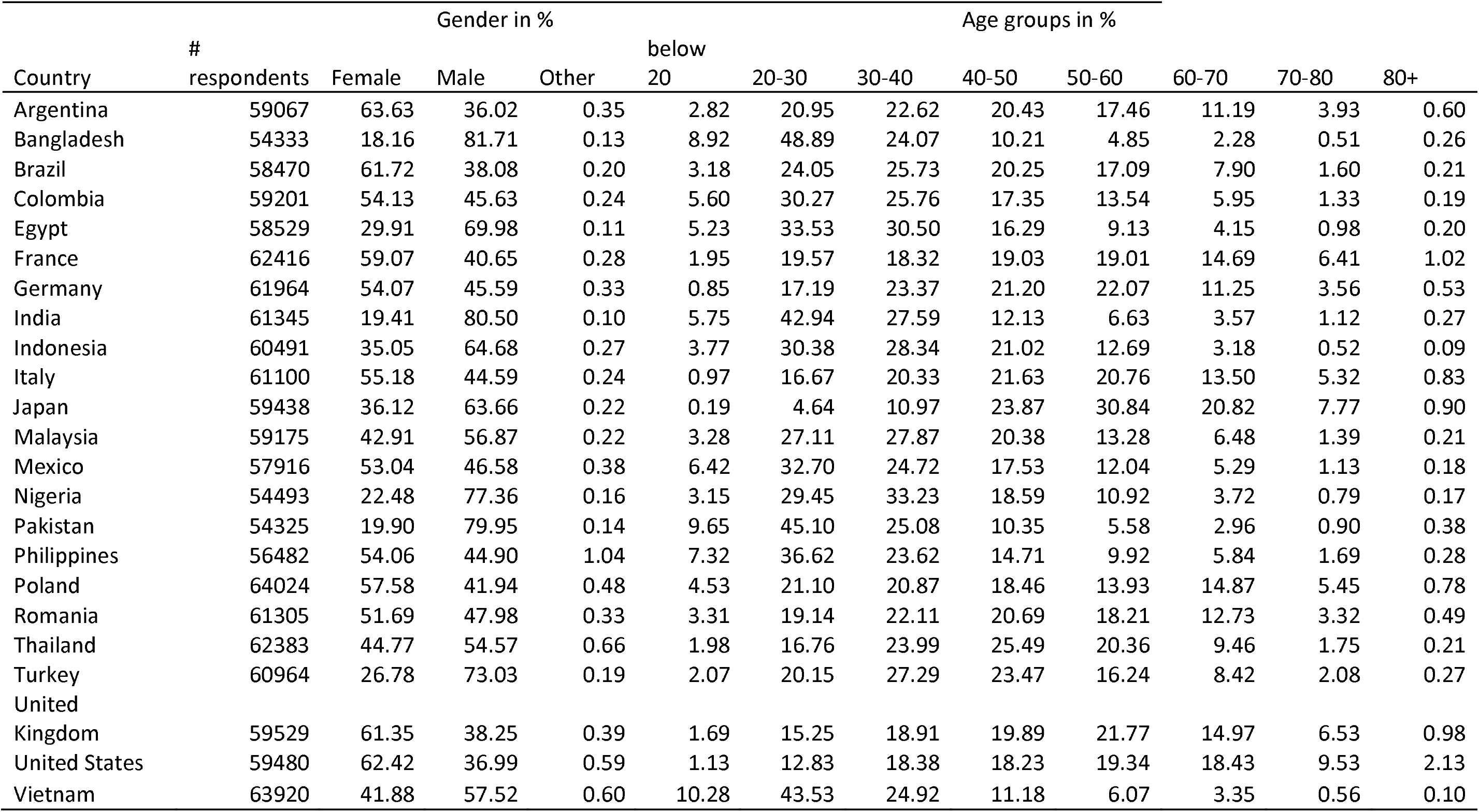
Country-level responses by gender and age groups.

### Measurement

The survey asked respondents whether they intend to get vaccinated against COVID-19 with possible responses of “Yes”, “No”, and “Don’t know.” Wave 15 to 18 (February – March 2021), additionally offered “I have been vaccinated” as an option, which for some analysis (see below) we coded as if respondents answered “yes” to their intent to vaccinate. Given the relatively small proportions of individuals receiving vaccinations at the time of survey, this coding choice does not appear to impact our substantive conclusions.

### Analytical Strategy

To understand variance in vaccine intent over time and space, we started by conducting Chi square tests to establish that vaccine acceptance significantly differed between country and over time (over all countries: χ^2^(66) = 107374, over time: χ^2^ (51) = 82132, p’s < 2.2.e-16). We then plotted and visually analyzed the distributions, with a plot of overall intent in the 23 countries over time for the 18 two-week intervals. Finally, we examined quarterly temporal changes across United Nations M49 geo-scheme, which largely reflect continents ^15^. We used the R software environment for all steps of the data analysis ^16^.

## Results

Answers were pooled for each country over all 18 waves, with more respondents willing to get vaccinated than were not (*M*_Global intent_ = 63.1%, *SD* _Global intent_ = 10.9%). In other words, a majority of respondents in the 23 sampled countries were willing to get vaccinated over the course of the pandemic. In the most recent wave of the survey (i.e., Wave 18), this proportion reached its highest surveyed level (*M*_Global intent_ = 72.7%, *SD* _Global intent_ = 10.8%).

Figure 1 shows that while no global longitudinal pattern exists, there is some visual indication that high-income countries in Europe (e.g., United Kingdom, Poland, Germany, France, and Italy) experienced increased vaccine intent over time, a pattern also seen in the United States. Notably, both the UK and France present a u-shaped pattern, while other European countries show early stability and a more recent increase. In other parts of the world, willingness to get vaccinated declined among respondents, particularly in middle and lower income countries (e.g., Colombia, Egypt, Indonesia, Malaysia, but see also Japan). In a third group of countries (e.g., Turkey, Vietnam, India), intent remained relatively stable among respondents.

**Figure 1:**
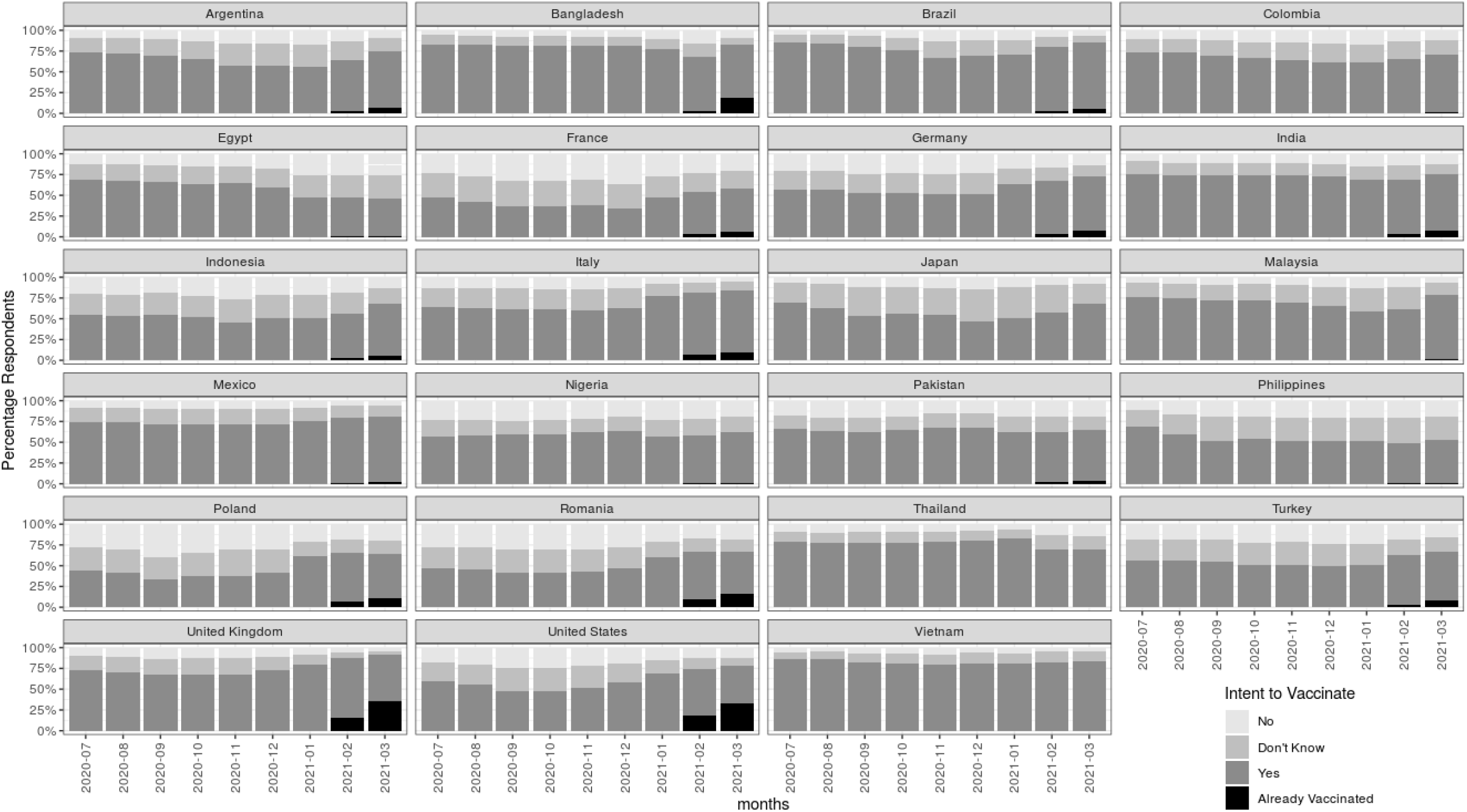
Intent to vaccinate per country and month (July 2020 – March 2021)

**Figure 2:**
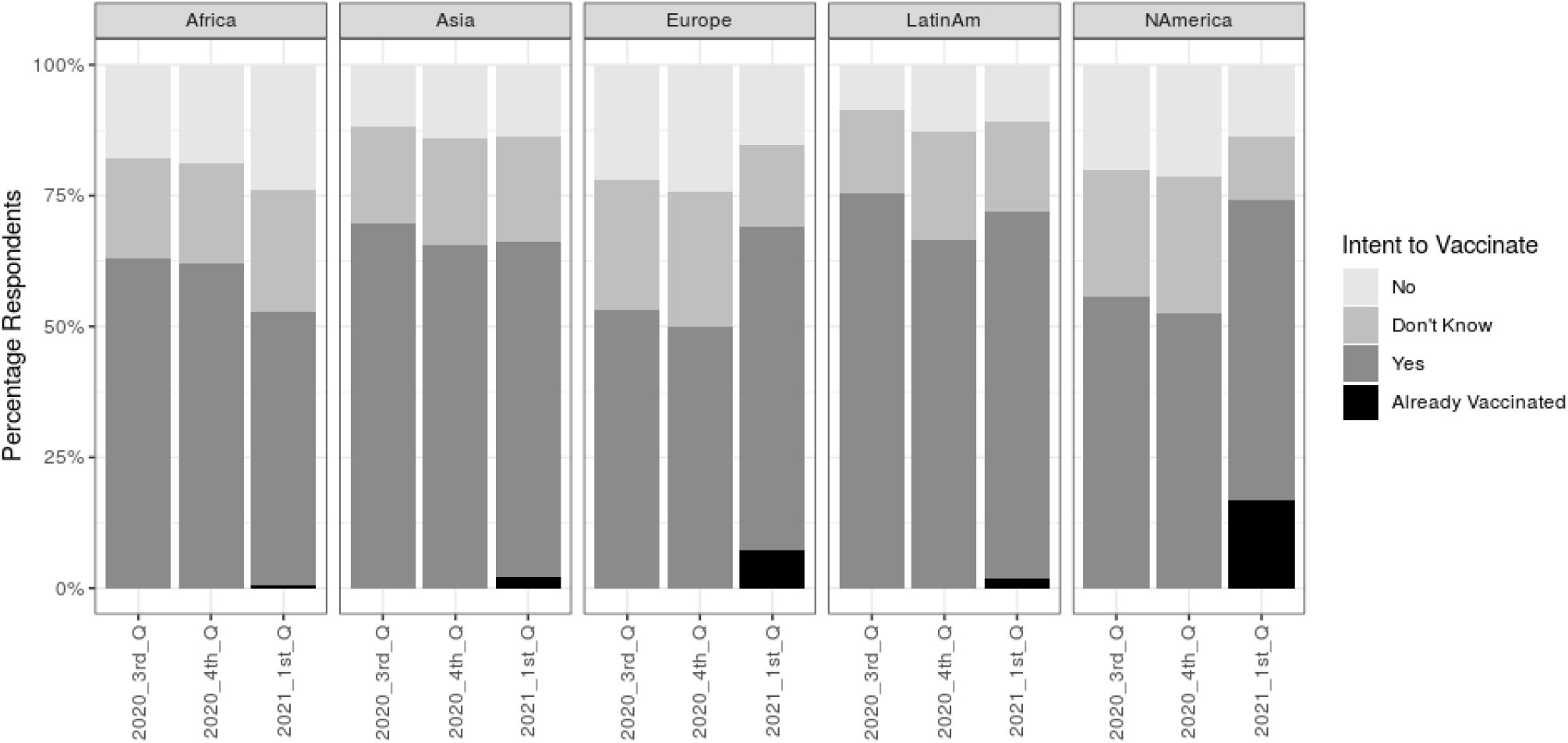
Intent to vaccinate in sample countries aggregated to UN region by quarter.

**Figure 3.**
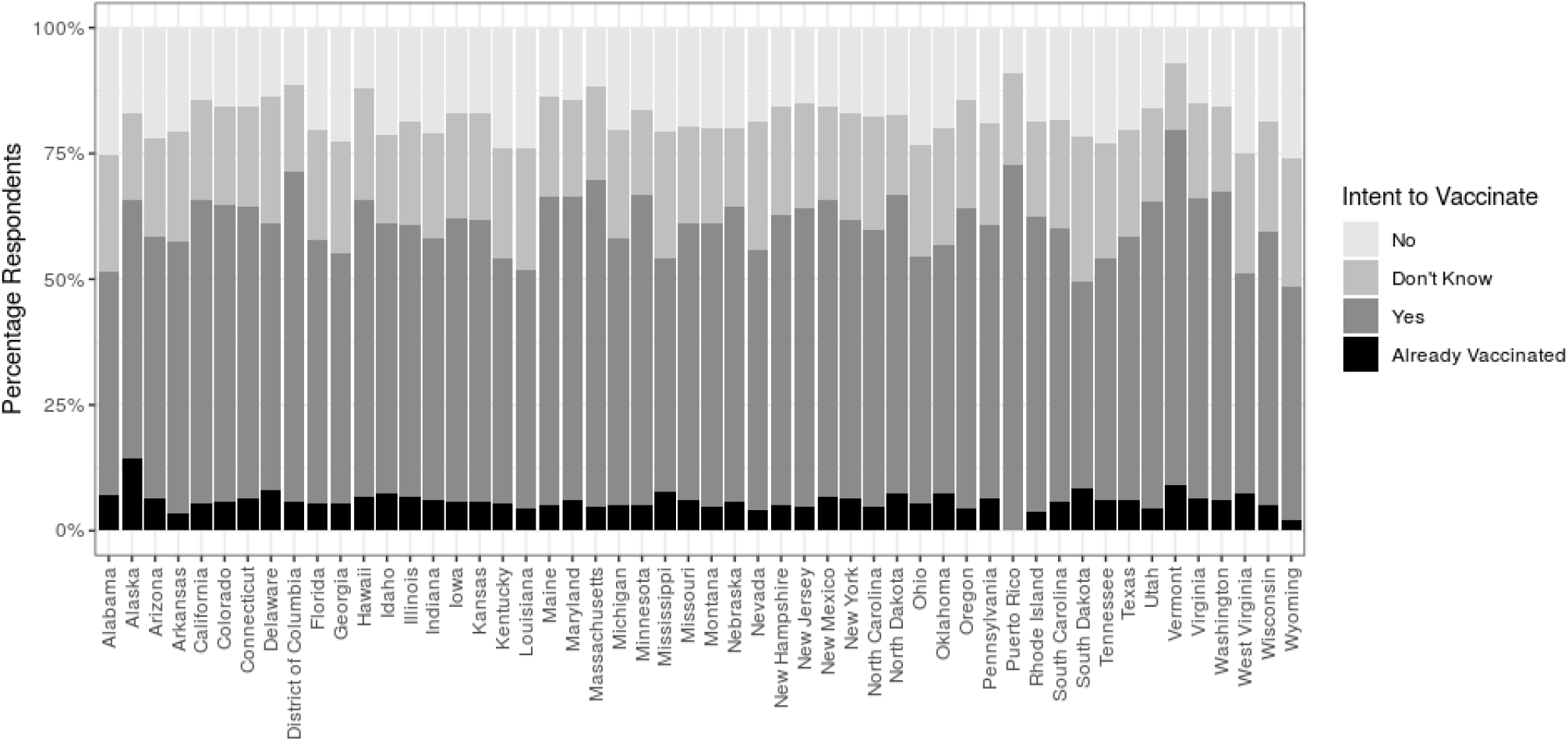
Intent to vaccinate across U.S. states.

To further examine temporal changes across geographic contexts, we aggregated countries to United Nation regions and waves to quarters. We find clear disparities in vaccine intent in the 3^rd^ quarter of 2020 with European and U.S. respondents expressing the lowest levels of intent. In all regions, willingness decreased slightly from the 3^rd^ to the 4^th^ quarter of 2020. Yet, among respondents from the U.S. and Europe, willingness rose sharply from the last quarter of 2020 to the first quarter 2021. While it remained largely stable in most Asian countries, it steadily decreased in one of our two African countries (Egypt).

This figure also illustrates the uneven roll-out of vaccines. A closer look at U.S. states also supports no vaccine hesitancy as rarely more than 25 percent of respondents had no intention to get vaccinated.

## Discussion

Using a unique longitudinal survey with over one million respondents from 23 countries, we found that vaccine intent among our respondents was higher than non-intent in all countries. Indeed, the average intent to vaccinate across countries over time has recently reached an all-time high during the pandemic. Importantly, we identify clear increases in acceptance as countries increasingly deployed the vaccines on a larger scale – but not immediately after the first approvals of the vaccines. Beyond that, our data tentatively suggests that, since the start of vaccination, the proportion of undecidedness has declined, although sampling bias cannot be ruled out. Finally, we found that not all countries shared this optimism - with acceptance rates stagnating in some countries or in the case of Egypt, actually declining.

Our central finding is much more positive than earlier studies suggest^9-10^, and indeed we can see a clear upward trend in vaccine acceptance in most countries in our sample. Specifically, vaccine intent rose since the end of 2020 among respondents in high-income countries in Europe (e.g., United Kingdom, Poland, Germany, France, and Italy), as well as the United States; and - to a lesser degree - in some middle-income countries (e.g., Mexico, Argentina), since the end of 2020. That acceptance increased in the beginning of 2021 is notable since this marks the actual mass distribution of vaccines in many of the aforementioned countries. Unfortunately, vaccine intent has remained constant in other parts of the world, though some countries evidence high vaccine intent overall (e.g., Vietnam, 83.8%), with other countries remaining stable at much more moderate levels (e.g., Philippines, 58.8%). Among the 23 countries, only Egypt showed a clear and consistent decrease in intent to vaccinate over time (from 68.3% in July to 47.5% in March). Finally, the general trend suggests that the portion of undecided respondents declined more quickly than those who outright opposed vaccination, although both positions shrank towards the end of the year.

These results send a clear message to politicians. A large portion of citizens across the world is willing to get vaccinated - in most countries in our sample this is enough to reach conservative levels of herd immunity ^17^. As such, the key barrier to vaccination may not be vaccine hesitancy, but rather the distribution of vaccines. Prompt and equal distribution of vaccines as envisioned by the COVAX-strategy^18^ should, therefore, be a top priority. This is of utmost importance since gaps in global vaccination increase the likelihood that new geographic variants emerge that are able to escape immune reactions. Further, vaccine enforcement, as considered by some countries, might not be needed ^19^. As most countries deliver the vaccine for free, also economical barriers for individuals are absent.

The second call for action goes to the scientific community and health communicators alike. We argue that researchers should focus their attention on (a) factors that can help to explain actual vaccination and (b) pockets of skepticism in some countries that we observed despite overall high and increasing vaccine intent. Identifying barriers that may lead to an intention-behavior-gap,^20,21^ as well as elucidating the motives and specific make-up of people who oppose the COVID-19 vaccination will aid a global campaign for vaccination. In turn, this will allow public health practitioners to better allocate resources for promoting vaccination in the places and for the populations most at-risk for missing vaccines or outright rejecting vaccination. In this regard, more research is needed on macro-level factors that might explain differences between regions, countries, or states (e.g., human development indices, health system performance and distribution, history of vaccination). Beyond that, more research is needed to understand those pockets of vaccine hesitancy within countries by including socio-psychological and cultural factors (e.g., personality traits, information consumption) and COVID-19-specific factors (e.g., risk perceptions around the vaccine and the virus in general) that matter not only for the current pandemic, but for the public health crises that follow in the future, as well.

## Conclusion

To curb the COVID-19 pandemic, citizens globally need to get vaccinated. While mass vaccination poses manifold challenges in terms of distribution – particularly on a global level – a broad acceptance of the vaccine among the public is a necessary condition for success. We found that vaccine acceptance among our respondents was higher than non-acceptance in all countries. Temporally, our results indicate that most countries are showing a positive trend towards higher acceptance, though low stagnant rates and the decrease in acceptance in a few countries is alarming. As more vaccines become available, global health officials need to identify changes in public opinions about the vaccine, identify barriers to actual vaccination and communities that are least likely to accept vaccination. This will ensure that the public’s memory of the morbidity and mortality associated with COVID-19 does not entrench resistance to immunization.^22^

## Data Availability

Microdata is only available after signing DUA with MIT and Facebook, we therefore cannot provide those data.
Aggregated data can be found at https://covidsurvey.mit.edu/

## Footnotes

1 N total sample = 1,425,172; but not all provided age and gender, hence N demographics = 1,370,350)

